# The effects of urolithin A supplementation on muscle strength, muscle mass and physical performance in humans - a systematic review

**DOI:** 10.1101/2025.07.10.25331277

**Authors:** Philippa Watts, Claire McDonald, Avan A Sayer, Miles D Witham

## Abstract

**Background:** Urolithin A, a stimulator of mitophagy, has been proposed as a therapy to improve skeletal muscle function via its beneficial effect on mitochondrial health. We aimed to systematically review existing evidence of the effect of urolithin A on measures of muscle strength, muscle mass and physical performance in humans.

**Methods:** We conducted a systematic review according to a prespecified protocol. Databases (PubMed, EMBASE, CINAHL, Scopus, ISRCTN.com and Clinicaltrials.gov) were searched from inception to 31^st^ May 2025, with hits screened by two reviewers. We included randomised controlled trials comparing urolithin A to placebo or usual care. We sought outcomes measuring muscle strength, muscle mass or physical performance. Risk of bias was assessed using the Cochrane Risk of Bias 2 tool. Results were grouped by outcome type and where applicable pooled using random-effects meta-analysis.

**Results:** We identified 194 titles, of which three studies were eligible for inclusion. Included studies recruited 174 participants, and mean age ranged from 24 to 72 years. All studies were placebo controlled and examined doses of 500mg/d or 1000mg/d of Urolithin A. Overall risk of bias of included studies was low; urolithin was well tolerated with good adherence to therapy. Four of 12 included outcome measures showed a statistically significant positive effect of urolithin A on muscle strength and physical performance, with a further seven outcomes demonstrating non-significant improvements in muscle strength or physical performance. Pooled analysis of six-minute walk distance from two trials showed a non-significant improvement in walk distance with urolithin A (23m [95% CI −6m to 52m, p=0.12, I^2^=0%]). One study measured muscle mass but found no improvement in mass with urolithin supplementation.

**Conclusions:** Insufficient evidence exists to support the use of urolithin A to improve muscle function in any population at present, but existing data support conducting larger randomised controlled trials in a range of target populations.

## Introduction

Enhancing skeletal muscle function is crucial not only for physical performance but also for maintaining overall health. This is especially true for older adults, in whom age-related muscle loss and weakness (sarcopenia) is a major health challenge [1,2]. Sarcopenia is an important contributor to frailty, disability and dependency in older adults [3]. Muscle weakness is also of relevance to recovery after severe illness or major surgery and is a common accompaniment to many chronic health conditions [4]. Resistance exercise is currently the only intervention consistently shown to improve muscle mass and function in individuals with sarcopenia [5]; however, many people are unable or unwilling to engage in the level of exercise required to produce meaningful improvements [6,7]. There is therefore an urgent unmet need for novel therapeutic strategies that can improve muscle strength and mass, both for people with sarcopenia [3], but also to enhance recovery from acute illness.

The determinants of muscle strength and mass, and of the pathophysiology of sarcopenia, are complex and multifactorial [8]. Among the various mechanisms implicated, mitochondrial dysfunction has emerged as a central pathogenic contributor. Mitochondria play a critical role in numerous cellular processes, including ATP and energy production, regulation of intracellular calcium homeostasis, modulation of cell proliferation, and integration of apoptotic signalling pathways [9]. Dysregulation of mitochondrial quality control mechanisms results in the loss of mitochondrial integrity, precipitating cellular dysfunction and muscle degeneration. Importantly, the accumulation of damaged mitochondria has been demonstrated to trigger motor neuron and muscle fibre death, thereby underscoring their pivotal role in the onset and progression of sarcopenia [10].

Urolithin A (UA) is a gut microbiome–derived postbiotic metabolite produced from ellagic acid, a polyphenol found in foods such as pomegranates, berries, and walnuts [11]. Preclinical studies have shown that UA can stimulate mitophagy—the selective degradation of dysfunctional mitochondria—thereby promoting mitochondrial health [12]. Studies in nematodes and both young and aged rodents have demonstrated that Urolithin A improves muscle strength and increases endurance [13].

Importantly, from a clinical translational perspective, Urolithin A has been shown to be safe, well-tolerated, and bioavailable in humans [14]. Emerging clinical evidence suggests that it may offer health benefits in age-related conditions such as cardiovascular disease, neurodegenerative disorders, and osteoarthritis [12]. Given its mitochondrial-targeting mechanism of action, Urolithin A has the potential to enhance muscle function and physical performance by improving mitochondrial quality control in skeletal muscle—making it a compelling candidate for enhancing muscle function.

To date, no systematic review has been conducted to evaluate the effects of Urolithin A supplementation on muscle mass, strength, or physical performance in human clinical trials. Such a review is critical to consolidating the current evidence, identifying gaps in the literature, and guiding future research and clinical application. The objective of this study was therefore to systematically review the evidence that Urolithin A supplementation can improve measures of muscle strength, physical performance and muscle mass in humans.

## Methods

We performed a systematic review of randomised controlled trials in humans. We used a prespecified protocol, which was registered on Open Science Framework prior to conducting the review. The protocol can be accessed at: https://osf.io/kzvyu.

We included randomised controlled trials comparing urolithin A with either placebo or usual care that enrolled human participants aged >=18 years in any setting. Studies examining healthy individuals, people with impaired physical performance, or people with specific conditions (including sarcopenia) were all eligible for inclusion. Studies had to include at least one measure of muscle strength, physical performance or muscle mass. No limits were placed on the duration of intervention or follow-up. Trials with identical co-interventions in each arm were eligible for inclusion. We excluded trials where a full-text report (journal paper or registry results summary) was not available. We also excluded trials where urolithin A was used with a co-intervention in the treatment arm on (e.g. urolithin plus exercise, vs usual care). We excluded observational studies, non-randomised trials or trials without a control arm, and studies not using purified urolithin A supplementation (i.e. those using unpurified food extracts).

We searched the following databases: PubMed, CINAHL, EMBASE, Scopus, along with the ISRCTN.com and Clinicaltrials.gov trial registries. Searches were run from the earliest date in the database to the 31^st^ of May 2025. No limits were placed on searches. Search terms used are given in Appendix A. Search results from each database were deduplicated in EndNote and then loaded into Rayyan for analysis. Titles were then screened independently by two reviewers (PW and MDW); studies were excluded only if both reviewers agreed that the title was not relevant. The process was then repeated for abstracts of those studies remaining in the selection process. Full-text studies were screened by both reviewers independently with final decisions on inclusion made by consensus.

Data items from each included study were extracted by one reviewer (MDW) and checked by a second reviewer (PW). We sought baseline study information including age, sex and description of the included population, intervention dose, duration and comparator. Muscle strength measures sought included, but were not limited to handgrip, quadriceps strength, or any other test measuring the strength of a muscle group or a compound movement. We sought strength-based physical performance measures including, but not limited to timed up and go test, Short Physical Performance Battery, sit to stand test, short walk speed. Physical performance measures of endurance included but were not limited to: 6-minute walk, 12-minute walk, cycling time, VO_2_ max (by treadmill or cycle), incremental shuttle walk test, seated step test, arm curl test, push-off force, recovery heart rate, or treadmill endurance time.

### Risk of bias assessment

Risk of bias for each included study was evaluated using the Cochrane Risk of Bias 2 tool [15]. Two reviewers (PW and MDW) independently evaluated risk of bias, with differences resolved by consensus. No quality threshold was applied to the inclusion of studies in the review.

### Data synthesis

Descriptive data on study characteristics and main outcomes were summarised in tabular form, with outcomes grouped as muscle strength outcomes, endurance exercise outcomes and muscle mass outcomes. Where incomplete data on the magnitude of treatment effects was reported (e.g. p values only, missing measures of dispersion), the missing data were calculated from available published data where possible, to give mean difference between groups with 95% confidence intervals [16]. Where dispersions and point estimates were reported only in graphical form, Image-J software [17] was used to measure these values on graphs to derive missing values. Meta-analysis of between-group change in outcomes was performed using RevMan software (v5.3) using weighted squares method with random effects models.

## Results

After deduplication, the search identified 194 titles. After review of full-text papers, three studies were included in the final analysis [18-20]. Details of the screening process are given in the PRISMA flow diagram (Figure 1). All included studies were parallel-group, placebo-controlled, randomised trials, and a total of 174 individuals were randomised across the three studies. The mean age ranged from 24 to 72 years, with diverse populations – young strength-training athletes, middle-aged individuals who were overweight or obese, and older people. No study specifically targeted older people with sarcopenia. Intervention duration ranged from 8 weeks to 4 months, with a dose of 1g/day used in all studies. One trial [19] also included a dose of 500mg/day. Adherence to study medication was reported as being high for all three studies, and dropout rates for participants were low (<10%) in all studies, with no excess of adverse events reported for participants taking Urolithin A in any of the included studies.

**Figure 1:**
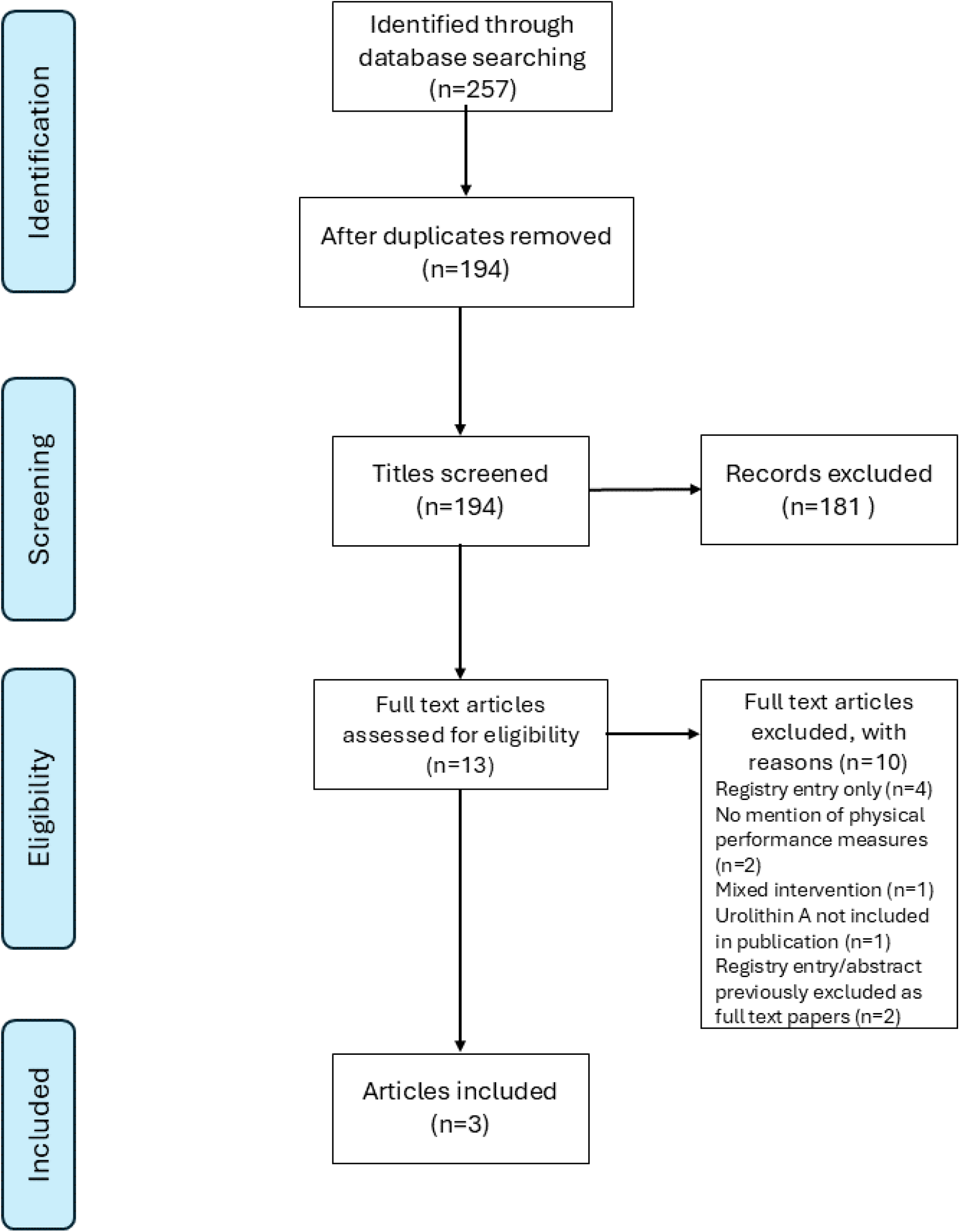
PRISMA flow diagram.

### Risk of bias assessment

Results from the RoB2 tool are given in Figure 2. Two trials [18,19] had an overall low risk of bias; one trial [20] had some concerns due to a lack of reported details regarding the randomisation and allocation concealment process, as well as insufficient details about the statistical analysis plan.

**Figure 2:**
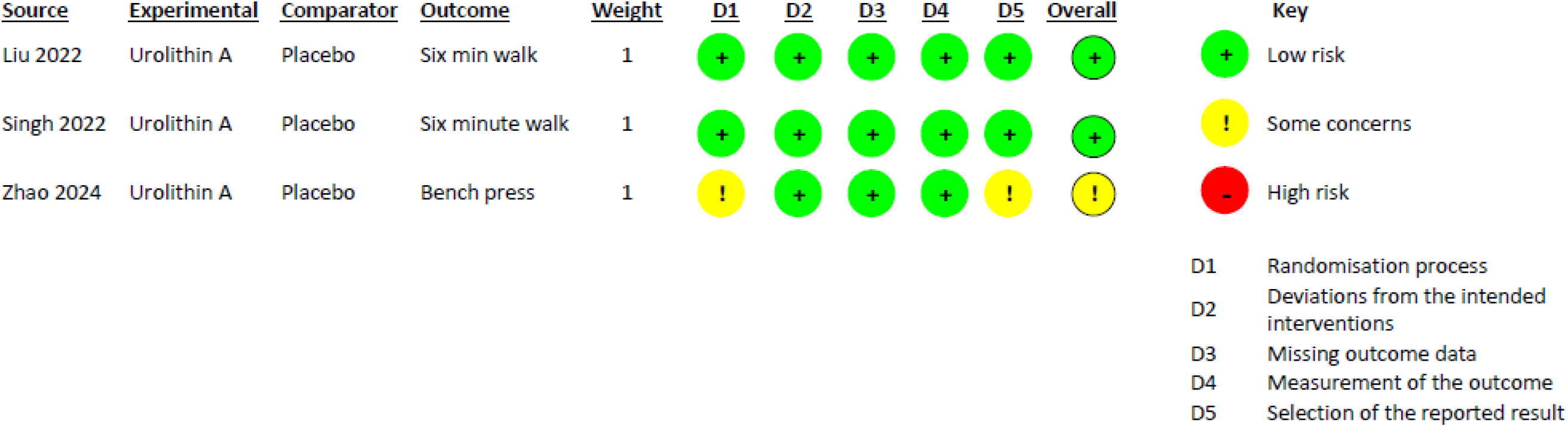
Risk of Bias assessment.

### Main results

Main outcomes from the three included trials are summarised in Table 2. Due to the complex analysis undertaken to derive significance values in Zhao et al [20] (including multiple-testing adjustments), the dispersion and significance values in the published article could not be reconciled. We therefore recalculated the 95% confidence intervals using the mean of standard deviation measures derived from graphs in this publication.

**Table 1:** Details of included studies.

| *Author/*<br>*year* | *N* | *Population* | *Mean*<br>*age*<br>*(years)* | *Female*<br>*%* | *Dose* | *Comparator* | *Duration* | *Outcome measures* |
| --- | --- | --- | --- | --- | --- | --- | --- | --- |
| Liu<br>2022<br>[18] | 66 | Older adults (age 65-90) | 72 | 76 | 1000mg/d | Placebo | 4 mths | 6 min walk distance<br>Forearm muscle contractions to fatigue |
| Singh<br>2022<br>[19] | 88 | Sedentary, overweight (BMI 25-35), middle-aged (40-65 yrs). No concomitant conditions requiring medication | 52 | 64 | 500mg/d,<br>1000mg/d | Placebo | 4 mths | Hamstring and quadriceps peak torque<br>Handgrip strength<br>6 min walk distance<br>Lean mass (DXA) |
| Zhao<br>2024<br>[20] | 20 | Young adult males undergoing resistance training | 24 | 0 | 1000mg/d | Placebo | 8 wks | Max bench press<br>Max squat<br>Max voluntary isometric contractions (quadriceps)<br>Repetitions to failure (bench press) |

DXA: Dual x-ray absorptiometry

**Table 2:** Outcome measures and effect sizes.

|  | Outcome | Study | Time point | Intervention |  |  | Placebo |  |  | Treatment effect as mean difference | P value |
| --- | --- | --- | --- | --- | --- | --- | --- | --- | --- | --- | --- |
|  |  |  |  | n | value | SD | n | value | SD |  |  |
| Muscle strength | Quads average peak torque | Singh [19] | 0 v 4 mths | 29 | +4.7% | NR | 30 | -2.5% | NR | +7.2% | NS |
|  | Hamstring average peak torque | Singh [19] | 0 v 4 mths | 29 | +9.8% | NR | 30 | -9.8% | NR | +19.6%<br>(2.0 to 37.2) | 0.029 |
|  | Handgrip strength | Singh [19] | 0 v 4 mths | 29 | +5.1% | NR | 30 | +2.4% | NR | +2.7%<br>(-0.3 to 5.7) | 0.08 |
|  | Max benchpress | Zhao [20] | 0 v 8 wks | 10 | +3.00kg | 0.17 | 10 | 0.5kg | NR | +3.50 (SD 0.79)<br>(-1.2 to 8.2)** | 0.46 |
|  | Max squat | Zhao [20] | 0 v 8 wks | 10 | +1.35kg | 2.73 | 10 | -1kg | NR | +2.55 (SD 1.36)<br>(-4.45 to 9.55)** | 0.71 |
|  | MVC quads | Zhao [20] | 0 v 8 wks | 10 | +36.10 NM | 0.62 | 10 | -6 NM | NR | +43.5 (SD 0.77)<br>(24.1 to 62.9)** | 0.048 |
| Physical performance | 6 minute walk distance | Liu [18] | 0 v 4 mths | 30 | 61m | 67 | 30 | 43m | 73 | +18m<br>(95% CI -18 to 54) | 0.32 |
|  |  | Singh [19] | 0 v 4 mths | 29 | +7.0%<br>(33.4m) | NR | 30 | -0.1%<br>(-0.5m) | NR | +34m<br>(95% CI -18 to 86) | 0.098 |
|  | VO<sub>2</sub>max | Singh [19] | 0 v 4 mths | 29 | +10.2% | NR | 30 | -1.1% | NR | +11.3%<br>(-0.5 to 23.1) | 0.06 |
|  | Contraction to fatigue | Liu [18] | 0 v 2 mths | 30 | 95s | 116 | 30 | 12s | 147 | +83s<br>(95% CI 14 to 151) | 0.018 |
|  | Bench press reps to failure | Zhao [20] | 0 v 8 wks | 10 | +2.00 reps | 0.56 | 10 | +0.1 | NR | +2.00 (SD 1.22)<br>(1.39 to 2.61)** | 0.011 |
| Muscle mass | Lean mass (DXA) | Singh [19] | 0 v 4 mths | 29 | -1.0% | NR | 30 | -0.7% | NR | -0.3% | NS |

DXA: Dual x-ray absorptiometry. MVC: Maximum voluntary contraction. VO<sub>2</sub>max: Maximal oxygen consumption. NR: Not reported. NS: Not significant (p>0.05)
\*For Singh et al [19], placebo vs 1000mg UA was compared in all cases. \*\*95% CI recalculated using SD's measured from Figure 3 of Zhao et al [20]
*Italics: calculated values (reported values in non-italicised script)*

Across all three studies, four of the 12 outcomes measured showed a statistically significant effect – maximal quadriceps contraction and bench press repetitions to failure in young strength-training athletes [20], maximal hamstring torque in middle-aged overweight people [19], and contraction to fatigue in older people [18]. In the trial comparing two different doses of Urolithin A (500mg/d and 1000mg/day) [19], almost all muscle strength and physical performance measures improved more with the 1000mg/day dose than with the 500mg/day dose; only the 1000mg/day comparisons are included in this analysis. A further seven outcomes suggested positive effects of urolithin A but did not reach statistical significance.

The six-minute walk distance was the only outcome measured by more than one trial, and the results of a pooled analysis of this outcome are shown in Figure 3: the pooled effect estimate by random-effects model was +23m (95% CI –6m to 52m, p=0.12, I^2^=0%). None of the other trial outcomes were sufficiently similar to be combined in a meta-analysis. Only one trial measured muscle mass (using DXA scanning); no significant difference in muscle mass was seen between the Urolithin A and placebo groups.

**Figure 3.**
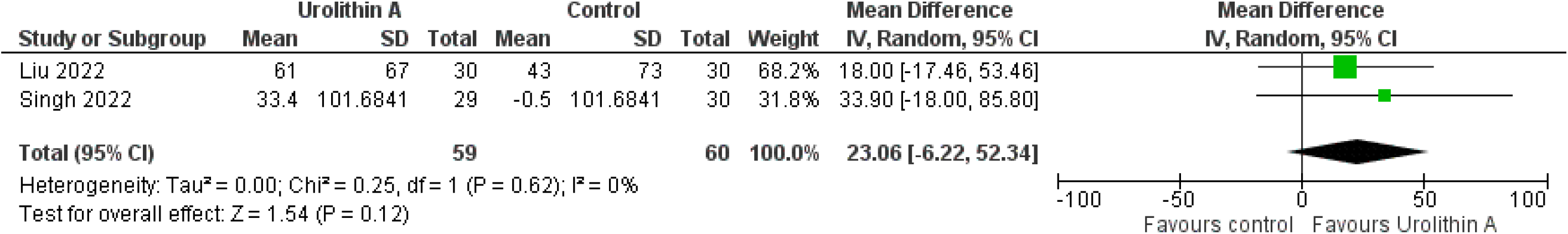
Pooled estimate of treatment effect for six-minute walk distance.

### Subgroup analyses

There were insufficient studies to perform pre-planned subgroup analysis by age or sarcopenia status. Similarly, insufficient studies were included for meaningful sensitivity analyses to be performed.

## Discussion

We found encouraging but limited evidence for clinically relevant effects of Urolithin A supplementation on measures of muscle strength and physical performance. Included trials were small in number and had insufficient statistical power to reliably detect the effect sizes found, but the included trials were overall at low risk of bias and analyses for outcomes tended to favour Urolithin A even when not reaching statistical significance. Only one trial included a measure of muscle mass [19], which showed no significant effect from Urolithin A.

The included evidence was limited by the small number of eligible trials, the heterogeneity of the studied populations, and the small sample sizes of the included trials. The overall body of evidence available for consideration in this review was therefore not sufficient to draw robust conclusions about the efficacy of Urolithin A either in general or in specific populations of interest. Limitations in the way that trials reported outcome data (in some cases, missing measures of dispersion, tabulated values for outcomes, or accurate measures of significance) required the derivation of missing data. Included trials were relatively short in duration – long enough to detect changes in muscle strength or physical performance, but less able to detect changes in muscle mass, although this was measured in only one trial. The small number of trials precluded more granular analysis (e.g. subgroup or sensitivity analyses, analyses of publication bias) and the wide range of outcomes measured precluded further meta-analysis.

We chose to conduct a rapid systematic review with a limited number of databases and a streamlined search process. It is possible that we did not therefore include all available trials but given the emerging nature of this field and the relatively small extant literature, the risks of this are low. The lack of limits on language and the independent screening and data extraction processes enhanced the robustness of the review. A limitation of our approach was that we did not seek data directly from authors; we do not think doing so would change the conclusions from the review, but given the limitations of some trial reports this should be considered for future, more extensive systematic reviews as the number of trials in this field grows.

Future reviews would benefit from access to individual participant data to enable recalculation of some outcomes, and this would also enable exploration by age, sex and baseline physical performance of groups most likely to benefit from Urolithin A supplementation. Most importantly though, larger trials of efficacy are required with sufficient sample sizes to reliably detect minimum clinically important differences for key measures of muscle strength and physical performance. Such trials need to be long enough to demonstrate efficacy over the medium term (several months of treatment) and long enough to detect any potential improvements in muscle mass. Trials targeting specific populations of particular clinical importance (e.g. older people with sarcopenia, people recovering from acute illness or surgery) are needed; such trials could also usefully explore the benefits of Urolithin A both in people who are undertaking exercise training as well as those who are not undertaking exercise.

In conclusion, these results suggest that Urolithin A cannot at present be recommended as a therapy to improve muscle strength, muscle mass or physical performance, but sufficiently encouraging signals are present to merit larger efficacy trials, particularly in populations with muscle weakness such as older people with sarcopenia and those recovering from illness.

## Data Availability

All data produced in the present work are contained in the manuscript

https://jamanetwork.com/journals/jamanetworkopen/fullarticle/2788244

https://www.cell.com/cell…/fulltext/S2666-3791%2822%2900158-

https://www.tandfonline.com/doi/full/10.1080/15502783.2024.2419388

## Acknowledgements

All authors acknowledge support from the NIHR Newcastle Biomedical Research Centre (reference: NIHR203309). CM, MDW and AAS acknowledge support from the NIHR Multiple Long-term Conditions Cross-NIHR Collaboration. MDW acknowledges support from the NIHR Newcastle Clinical Research Facility. PW acknowledges support from Newcastle Clinical Trials Unit.

## Appendix A Search terms used

(Randomi*ed controlled trial OR Controlled clinical trial OR Placebo OR Intervention OR Trial* OR Experimental medicine)

AND

Urolithin*

AND

(Muscle OR Sarcopenia OR Strength OR Physical performance OR Mitochondria* OR

Mass)

